# Body Weight and Cardio-respiratory Fitness: Predictors of Physical Function Capacity among Older Adults

**DOI:** 10.1101/2023.06.05.23291006

**Authors:** Eric A. Aloko, Munkaila Seibu, Daniel Apaak, Emmanuel O. Sarpong, Stephen R. Sorkpor, Edward W. Ansah

## Abstract

*Ageing is an inevitable part of human life, thus, everyone may grow and become old. The ageing process is characterized by reduced physical activity, accumulation of fat and loss of muscle mass resulting in weight gain and reduced cardio-respiratory function that leads to loss of physical function capacity. Therefore, the purpose of this study was to determine the extent to which body weight and cardio-respiratory endurance predict physical functional capacity of older adults in Navrongo, Ghana. This study employed quantitative cross-sectional design, using a multistage sampling method with 998 aged (60yrs+) participants. The senior fitness test battery, international physical activity questionnaire (IPAQ) short form for elderly, weighing scale and tape measure were used to collect the data. The independent t-test and multiple regression were applied to analyse the data. The results indicated that physical activity levels were generally minimal among both men and women with 53% of the population having MET of <600, but women were more overweight (BMI=28kg/m^2^), had reduced cardio-respiratory function* (M = 53.22, SD = 12.51) compared with men (M = 70.87, SD = 15.60) *and physical function capacity (<100 functional score). The results also indicate that body mass index and cardio-respiratory endurance are significant predictors of physical function capacity among older adults* F (2, 995) = 956.63, p < .05, R = 0.81, R^2^ = .658 and adjusted R = .657. *It is concluded that, body mass index and cardio-respiratory endurance are significant predictors of physical function capacity among older adults in Navrongo. It is recommended that, education on the health benefits is provided and regular participation in physical activity is done to promote regular exercise among these older adults. A longitudinal study is needed to explore the moderation-mediation role of physical activity on the relation of cardio-respiratory function and physical function capacity among older adults*.

## Introduction

Ageing is an inevitable for human life, and everyone will grow old. This process of ageing comes with limitations, including limitations in physical functional abilities (WHO, 2019). This becomes more apparent with the increase in body weight and compromised cardio-respiratory function. According to Magyari et al. (2018), body composition and cardio-respiratory endurance are two most studied physical fitness components that affect physical functional capacity of older adults. These authors explained physical functional capacity as person’s ease of performing physical and motor activities of daily living. Meanwhile, physical activity, exercise, rest, and nutrition are key elements that must be taken seriously by individuals irrespective of age to promote physical function capacity. Exercise is a systematic, repetitive body movement with the goal of enhancing or maintaining fitness, as opposed to physical activity-any movement of the skeletal muscles that results in energy expenditure (Garber et al., 2011). Thus, physical activity and exercise, nutrition, and rest are some of the determinants of body composition and cardio-respiratory endurance, that help an individual to transit into healthy aging (Magyari et al., 2018).

Ageing is a process that can be healthy or unhealthy, and characterized by decline in cardiovascular function (Bolton, & Rajkumar, 2019), increased blood pressure, compromised body composition, decreased muscle mass (sarcopenia), increased adiposity, and muscle fat infiltration, all leading to a decrease in functional capacity of the individual (Carbone et al., 2017). The age-related decline in physical function capacity resulting from reduced mobility and the increased efforts needed to perform daily activities, could ultimately lead to avoidance of meaningful physical activity and exercise, resulting in sedentary living. This predisposes the individual to cardiovascular diseases (CVDs), obesity, and other forms of disabilities (Magyari et al., 2018).

The ageing bracket is increasing in all continents of the world. For instance, the United Nation Department of Economic and Social Affairs, Population Division (2019) reported that there were 703 million people age 65 years or older globally in the year 2019. Accordingly, the figure is expected to more than doubled, reaching 1.5 billion by 2050. In Northern America and Western Asia, the number of elderly people is projected to climb at the quickest rate, from 29 million in 2019 to 96 million in 2050 (an increase of 226%). The second fastest rise in the number of older persons is foreseen in “sub-Saharan Africa, which is expected to grow from 32 million in 2019 to 101 million in 2050 [218%]” (United Nation Department of Economic and Social Affairs, Population Division, 2019, p.5).

Unfortunately, the ageing population is likely to increase the burden of chronic illness in all economies globally. According to the United Nations Population Division (2019), it is envisaged that the number of older adults with non-communicable diseases will increase too, with more disabilities and decline in physical functional abilities globally, with CVDs as part of the three leading causes of deaths. The case in low-and-middle-income countries (LMIC) is worse, as WHO (2019) revealed that over 80 percent of deaths relating to CVDs occurred in these countries including Ghana.

Sub-Saharan Africa (SSA) has a population rapidly reaching one billion (United Nations [UN], 2017). In addition, the UN estimated that 4.9 percent of the total population of persons living in SSA as at 2015 were 60 years and over, a proportion projected to reach 7.6 percent by 2050. Unlike most western countries where there is supportive built environment for exercise, SSA’s built environment is not exercise friendly, thereby increasing the avoidance of physical activity and exercise among the aged in particular (Wendel-Vos et al., 2010). However, it is anticipated that more than 60% of the total population of older adults in SSA have at least one or more physical functional limitations (Stuck et al., 2013). Therefore, governments in Africa and the African Union (AU) need to pay attention to this rise in the number of older adults and make conscious efforts to put interventions that will save and preserve their lives (Wendel-Vos et al., 2010). Among such interventions could be providing exercise education and creating exercise friendly environments where people can exercise safely outdoors with no challenges.

The number of people 60 years and over in Ghana from 2000 to 2010 has reduced from 7.2 percent to 6.5 percent (GPHC, 2010). Despite this reduction, it is projected that the number of older adults will triple by 2030 (Ghana Statistical Service [GSS], 2014). According to the GSS District Analytic Report for Kassena Nankana, the number of older adults in the Municipal capital as at 2010 was 9,728 and this number is projected to reach 20,000 by the next census (GSS, 2014).

A longitudinal study by the Navrongo Health Research Center (NHRC, 1998) using a self-reported measure revealed health and functional limitations among older people in Kassena-Nankana District. The study revealed that over 80% of the participants self-rated their health and physical function as good, which was to the manual farming activities of the majority of the people. Unfortunately, a follow up study in 2010 showed a different result, as over 85% of the participants self-reported at least two or more physical functional limitations (NHRC, 2010). The researchers however, were of the view that, the change in activities (abandonment of farming and other physically active activities to less physical activity ventures like trading) among the people contributed to the later results. Thus, these findings coupled with the rise in number of older adults calls for innovative interventions to promote physical activity and overall physical function capacity and health of these older people. However, there seem to be no sufficient evidence to back such physical activity and exercise intervention programmes. Therefore, the purpose of this study was to determine the extent to which body weight and cardio-respiratory endurance predict physical function capacity of older adults in Navrongo, Ghana.

## Materials and Methods

This study employed quantitative cross-sectional design to investigate the role of physical activity, body composition and cardio-respiratory endurance and their influence on physical function capacity of the older adults. The population included adults 60 years and older who live in Navrongo. Unfortunately, the Ghana Statistical Service’s 2021 National Population Census report does not yet include age distribution and the district level analyses are also not available. As a result, this study relied on the 2010 Population and Housing Census data, which provided that there were 1,663 senior adults in Navrongo with 941 women and 722 men (GSS, 2010).

Navrongo has been put into four cardinal zones namely, North, South, East and West by the Navrongo Health Research Center (NHRC, 2012). Using a multistage sampling approach, 998 older adults participated in this study. Firstly, using the quota sampling, 60% of males and females were sampled, giving a sample size of 433 males and 565 females.

The North, South, East and West comprises 7, 9, 7 and 6 suburbs, respectively. The sample sizes (433 males and 565 females) were divided into four and assigned to each suburb, making it 108 males and 141 females each for the following zones (North, East and West) except for South which comprised 109 males and 142 females. Five research assistants were assigned to each cardinal zone for the data. The research assistants visited the various households in each suburb of the various cardinal zones for data collection. The convenience sampling was used to include participants who were available at the time of visit to each household and were willing to participate in the study. Participants who were readily available at home and were willing to participate in the study were made to sign or thumbprint a consent form after the purpose of the study was explained to them. Those who could read and write signed, but for those who could not read nor write, the content of the consent form was explained to them in their local dialect (Kassem and Nanakani) and those who agreed to take part thump printed the consent form.

For purposes of anonymity and confidentiality, information that identifies the participants were not obtained. At the end, 108 males and 141 females were conveniently sampled from North, East and West zones while 109 males and 142 females were conveniently sampled from the south zone.

### Setting

The Kasena Nankana Municipal is one of Ghana’s 260 Metropolitan, Municipal and District Assemblies (MMDAs) and is a member of the Upper East Region’s fifteen (15) Municipalities and Districts, with Navrongo serving as the regional administrative center. Within the Guinea Savannah woods is Navrongo. Between latitudes 11o10’ and 10o3’ North and longitude 10o1’ West is about where it is situated. Navrongo is the administrative center. The town shares boundaries with Paga and Bolgatanga, Sandema. However, it is also bounded to the north of the Ghana by Burkina Faso. The number of household in Navrongo stood at 32,000 with a total population 109, 944 with 53,676 males and 56,268 females (Ghana Statistical Service, 2013). The main occupation of the people in the Municipality is farming and trading. The built environment of the Municipality is not exercise friendly. Also, there exist a poor culture towards meaningful physical activity and exercise in the Municipality. Hence, sedentary living is gradually becoming an order of the day in the Kassena Nankana Municipality.

### Instruments

This study adopted four data collection instruments, namely, the Senior Fitness (Functional Fitness Test) battery, Omron weighing scale, tape measure, and international physical activity questionnaire (IPAQ) Elderly short form. Rikli and Jones (2013) created the Senor Fitness Test battery as part of Fullerton University’s lifelong wellness programme, as it is frequently called the Fullerton Functional Test. It is a straightforward, user-friendly battery of tests that evaluates older individuals’ functional fitness, like the aerobic fitness, strength and flexibility among adults, using minimal and inexpensive equipment. Within the test battery are specific tests and their reliability coefficients; “chair stand test (.88), arm curl test (.79), sit and reach test (.81), back scratch test (.82), up and go test (.84) and 2-minute step up test (.81)” and a composite reliability of .83 for PFC (Rikli, & Jones, 2013). Reliability coefficients estimated from current data were .79, .80, .77, .78, .80, .79 for chair stand test, arm curl test, sit and reach test, back scratch test, up and go test and 2-minute step up test respectively and a general reliability of .81 for physical functioning capacity, respectively.

The Omeron scale and meter tape were used to obtain the weight and height of the participants. Vasold et al. (2019) reported a reliability of .91 for the weighing scale and .90 for the meter tape. Data from the current study also yielded reliability coefficient of .88 and .91 for the weighing scale and meter tape, respectively. We used the IPAQ-Elderly short form to measure the physical activity levels of older adults, with reliability and validity values of .81 and .84 when tested across six centers in 12 countries (Craig et al., 2003). Moreover, the current study yielded a reliability of .79.

### Procedure

The Institutional Review Board of University of Cape Coast granted the ethical clearance (ID-UCCIRB/CES/2022/30). To collect the data, permission was sought from household heads and the purpose of the study well explained to them. Participants were also assured of confidentiality of their information. Also, for purposes of anonymity, names of participants names were not taken. For inclusion criteria, only older adults who could walk and raise their upper limbs were considered for this study.

First aiders were also on standby during the fitness test to take care of any eventualities. Data was collected from participants at their homes. Physical function capacity for each participant was assessed through the five physical functional test batteries. To assess physical activity levels, participants completed the IPAQ elderly short form which asked asked them to indicate the number of days in the week they walked, did moderate and vigorous activities and the amount of time spent doing such activities. Cardio-respiratory endurance was measured using 2-minutes step-up test. A 2-minutes test is performed by making a mark of the corresponding mid-point between the iliac crest and the knee on a wall. A participant stand with the should to wall and on the command by the instructor, the participant lifts (step-up) the knees one after the other at the level of the mark on the wall as fast as he/she can. The total number of step-ups taken within 2-minutes are recorded for each participant. Body composition was assessed using body weight and height to compute for their BMI. Body weight was measured to the nearest 0.01km using the omron weighing scale where participants wear a light cloth and without footwear. Height was measured using a meter tape attached to a wall. Each participant (without a footwear) stood with the back facing the wall and heels touching the wall. A flat plastic meter rule is then place on the head of the participant in line with the meter tape. The corresponding number was then recorded in meters as the height of the participant. To obtain the BMI of each participant, the weight (kg) was divided by the square of the height in meter (Weight/Height^2^). The data were collected between August and December, 2022 between 8 AM and 4 PM during weekends when the participants were likely to be at home.

### Data Analysis

Data were entered into Statistical Package for Social Science, IBM, version 25 for windows and went through screening process before statistical analysis. Demographic variable (gender) was analysed using frequency and percentages. Physical activity levels of older adults were analysed using frequency and percentages. Moreover, differences in gender by cardio-respiratory endurance and body composition were analysed using independent sampled t-test. Gender a categorical variable, body composition, cardio-respiratory endurance, physical function capacity and physical activity were measured on a continuous scale, thus, t-test analysis (Mishra, Singh, Pandey, Mishra, & Pandey, 2019). Finally, multiple linear regression was computed to predict physical functioning capacity from cardio-respiratory endurance and body composition values. The variables included a continuous dependent variable and two independent variables (Chatfield, 2018). Results were reported on correlation, R, R^2^, ANOVA analysis (regression df and residual df, F-ratio and p-value) and the coefficients (beta-values and p-values).

## Results

Demographic characteristics of participants was obtained using frequency and percentages

Table 1 above showed that, there were 565 males representing 56.6% and 433 females representing 43.4%.

**Table 1:** Data showing number of males and females in the study.

| Variable | Frequency | Percentage |
| --- | --- | --- |
| Male | 565 | 56.6% |
| Female | 433 | 43.4% |

To determine the physical activity levels of these older adults, frequency and percentages analysis were estimated, and the results are presented in Table 1. The results indicate that 470 (47%) participants were physically inactive while 528 (53%) were minimally active. Meanwhile, none of the older adults engaged in health-enhancing physical activity levels. According to the WHO (2019), minimally active levels of PA are not recommended for health promotion by individuals of all age categories. Therefore, since 47% were physically inactive and 53% being minimally active, the older adults in Navrongo were not meeting the WHO physical activity guidelines of 150 minutes of moderate-to-vigorous intensity of physical activity for at least three times in a week.

To determine the differences in cardio-respiratory endurance, body composition, and physical function capacity by gender, three different independent sample t-test analyses were calculated. The findings revealed a statistically significant difference in cardio-respiratory endurance between male and female, t = (812) = 19.27, p < .05, where women recorded a lower cardio-respiratory endurance (M = 53.22, SD = 12.51) compared with men (M = 70.87, SD = 15.60). Hence, women had poor cardio-respiratory fitness levels. Also, there was statistically significant difference in BMI by gender, t (996) = -26.00, p < .05, with women having a higher BMI (M = 27.85, SD = 4.14) than the men (M = 21.28, SD = 3.22). Hence, the females in this group are overweight and, on the brink, of crossing into to obesity class 1. Furthermore, the results showed a statistically significantly difference in physical function capacity by gender, t (833) = 21.92, p < .05, with higher functional capacity for men (M = 101.79, SD = 15.95), than the women (M = 81.02, SD = 13.25). Hence, the women in this group may suffer more functional limitations.

To predict physical function capacity from cardio-respiratory endurance and BMI, a multiple regression was performed. The results indicate statistically significantly prediction for physical function capacity, F (2, 995) = 956.63, p < .05, R = 0.81, R^2^ = .658 and adjusted R = .657 from cardio-respiratory endurance, β = .774, t (995) = 38.56, p < .05) and BMI, β = -.089, t (995) = - 4.44, p < .05). Therefore, cardio-respiratory endurance and BMI are good predictors of physical function capacity among older adults in Navrongo (see Table 2 for details).

**Table 2:** Physical Activity Levels of Older Adults in Navrongo.

| Met Range | Description | Frequency | Percentage |
| --- | --- | --- | --- |
| < 600 | Inactive | 470 | 47% |
| 600 – 1500 | Minimally active | 528 | 53% |
| > 1500 | HEPA | 0 | 0% |

**Table 3:** Predicting Physical Functional Capacity from Body Composition and Cardio-respiratory Fitness.

| **Variable** | **B** | **Beta** | **T** | **p** |
| --- | --- | --- | --- | --- |
| Constant | 47.349 | . | 17.830 | .000 |
| Cardio-respiratory fitness | .837 | .774 | 38.857 | .000 |
| Body mass index | -.327 | .089 | -4.443 | .000 |
| R | .811 |  |  |  |
| R<sup>2</sup> | .658 |  |  |  |
| Adjusted R | .657 |  |  |  |

**F-ratio = 956.63, df = (2, 995), p < .05**

## Discussions

The aim of this study was to quantitatively explore the role of physical activity, body composition and cardio-respiratory endurance and their influence on physical function capacity of the older adults in Navrongo, Ghana. The finding reveals that these older adults in Navrongo were not meeting the WHO PA guidelines for health promotion and healthy living, the situation predisposes these older adults to health challenges including limitations in their daily physical functions. This result is very worrying and alarming as no participants met health-enhancing levels of physical activity. Therefore, the population of adults in Navrongo may be predisposed to metabolic disorders, and other chronic non-communicable diseases such as cancer, diabetes, cerebrovascular, and cardiovascular diseases (WHO, 2019). Physical inactivity among these adults is attributed to a decline in physical activity participation probably because of age (Magyari et al., 2018). Additionally, there seems to be no conscious effort by these older adults to carry out daily strength exerting activities. McPhee et al. (2016) found that age-related declines in physical activity are caused by both social and physiological factors. The avoidance of meaningful physical activity among the elderly is also influenced by physical limitations, disease, and discomfort, as well as retirement coupled with a loss of interest in social activities (including physical activity).

Furthermore, McPhee et al. noted that despite being aware of the advantages of greater physical exercise, many older persons continue to be sedentary and engage in no or very little physical activity. The 53% who are minimally active may have been very active in their youthful days. This might have helped them to transit into minimal healthy active ageing. Thus, efforts at promoting youth physical activity are important for healthy ageing activeness. For instance, a person must perform moderate-to-vigorous physical activity for at least 30 to 60 minutes, three times a week to reach the health-enhancing physical activity levels (1500MET-3000MET) (WHO, 2019). This, requires extra physical efforts from older adults, which may lead to few or no adult meeting such levels of physical activity, as may be the case with the elderly population in Navrongo (Carbone, Lavie, & Arena, 2017). Therefore, there may be the need to study, re-calibrate and re-design health-enhancing exercise threshold for older adults in rural areas and developing countries.

Age-related health issues might also lead to a decline in social interaction, including participation in physical exercise (Bernard, 2019). This justification is consistent with the Disengagement Theory of Ageing (Cunning & Henry, 961), which contends that as one ages, their talents, particularly their capacity to interact with friends and family, decline. Consequently, in comparison to the youths, older adults increasingly sever their relationships to others in their society and become more physically inactive and lonely. However, physical activity, even if initiated later in life, might delay the onset of impairment in the elderly (Lafortune et al., 2016). Additionally, a preventive effect against disability among the elderly can be achieved by lifestyle risk factors like engaging in regular physical activity (Moody-Ayers, Mehta, Lindquist, Sands, & Covinsky). Moreover, adults in some neighborhood may participate in less physical activity due to a poor constructed neighbourhood environment. Unfortunately, the built environment in Ghana is not exercise-friendly (Wendel-Vos, Droomers, Kremers, Brug, & Van Lenthe, 2010).

The environment plays a crucial role in promoting physical activity. For instance, WHO (2019) made the case that incorporating physical exercise into daily life, such as walking and cycling may help people to meet recommended levels of physical activity. Thus, WHO calls for a supportive built environment. Beyond social and individual factors influencing physical activity (Sullivan, & Lachman, 2017), evidence further suggests that well-designed exercise friendly communities are important (Smith et al., 2017). The importance of the built environment must be highlighted since it motivates people to engage in greater physical activity than a setting that is badly designed and does not support an active lifestyle (Smith et al., 2017). Perhaps, such as well-designed walkable path, recreational parks, cycling paths among others may increase physical activity which turn to lower the risk of CVDs, obesity, and other disabilities leading improve physical functioning capacity (Magyari et al., 2018).

The findings indicate that the elderly women have low cardio-respiratory function. Older people have substantially higher frequency of CVDs than the general population, but with gender differentiations. For instance, Malmborg et al. (2021) state that 77.8% of females and 70.8% of males ages 65 and 74 were diagnosed of hypertension between 2013 and 2017. Furthermore, the rates of diagnosed hypertension rose sharply, reaching 80.0% for men and 85.6% for women over age 75. Additionally, among older adults, obesity, gender, and age are particularly linked to hypertension (Hajar, 2016), which disproportionately affect women.

Elderly women in Navrongo having low physical activity levels and increase overweight may lead to more hypertensive cases (Hands et al., 2016). This may be due to lower stroke volumes of women and the slower increase in stroke volumes from rest to exercise which are also the most constant gender differences in the cardiovascular responses to endurance or "dynamic" exercise. Women’s lower peak oxygen consumption (VO_2_peak) and smaller stroke volumes at rest and during exercise may be related to gender-specific variations in heart size/mass and blood volume. Moreover, Jakovljevic (2018) confirms that the lower heart size of females compared to males is another morphological trait that appears to affect VO_2_ max. Elderly women may need extra attention to promote their functional capacity.

Our findings on gender difference in BMI also revealed that the female older adults are overweight and on the borderline of obesity class 1. This condition predisposes the women to the higher risk of non-communicable diseases such as coronary heart disease, strokes, and hypertension among others. Dogra (2017) claims that because there are disparities in BMI between the sexes, females are frequently regarded as having a higher risk of experiencing negative health effects and are designated as a high priority category for physical activity interventions. Additionally, increase in weight may result in excess body fat, and more cholesterol deposits in the blood, complicating the health of women by blocking the flow of blood to the brain, to-and-fro the heart and other organs, resulting in a compromised cardiovascular system (Elagizi et al., 2018). These are likely to increase the risk of hypertension and other chronic helath conditions among these women.

Furthermore, it is found that older women in Navrongo recorded a mean physical functioning capacity value of 81.02, below the average standard score of 100. This means that older women in Navrongo may suffer from physical functional limitations, because evidence (Poggiogalle et al., 2019) suggest that age plays a critical role in functional capacity (strength, endurance, agility, and flexibility), making daily activities more challenging especially women who are advanced in age. Therefore, when physical inactivity becomes prevalent among women, health-related fitness components, especially body composition and cardio-respiratory fitness are compromised. This can lead to a decline in physical function capacity, thereby, predisposing women to high risk of non-communicable diseases, orthopedic problems and other related illnesses.

It is also evident that BMI and cardio-respiratory endurance are good determinants of physical function capacity among these older adults. This finding reflects the statement by Magyari et al. (2018) that body composition and cardio-respiratory endurance are two health-related fitness components known to predict physical function capacity among older adults. Physical functioning capacity of individuals is key to their survival, but this could be compromised by being overweight or obese, having a poor cardiovascular fitness and other illnesses. It has been demonstrated that obesity in the elderly, as measured by a high BMI, is substantially associated with a deterioration in functional performance, which could result in disability. One possible explanation is that excess body weight can place additional strain on the joints and muscles, which can make it more difficult to perform physical activities. Additionally, carrying excess weight can increase the risk of developing chronic health conditions such as arthritis, diabetes, and CVDs, which further limit physical activity efforst and impair physical functioning ability of persons (Poggiogalle et al., 2019). As a result, older adults especially women are likely to suffer from physical functional limitations due to their high BMI values. Ponti et al. (2020) stated that the risk of disability and functional limitations should be taken into account together with individual’s history of being overweight or obese.

The link between cardiovascular endurance and physical functioning can be attributed to the fact that aerobic exercise, which improves cardiovascular endurance improve muscle strength, balance, and flexibility, all of which are important components of physical functioning ability (Žargi, Drobnič, Stražar, & Kacin, 2018). According to Connolly et al. (2017), engaging in regular aerobic exercise can reduce the risk of chronic diseases such as CVDs and diabetes, which can improve physical functioning capacity of persons.

The findings of this study have important implications for promoting healthy ageing. Thus, interventions that promote healthy weight management and cardiovascular fitness may help these older adults to maintain their physical independence and improve their overall quality of life (Munt et al., 2017). This may help reduce the burden of dependency rate, limits both personal, family and national healthcare cost, and minimise pressures on healthcare providers and the entire healthcare system of the nation.

## Limitations

Though this study provides very useful findings of physical activity levels, BMI and cardio-respiratory endurance and their relations with physical function capacity of older adults from rural community in a developing nation, there are some limitations. Other variables such as diet, medical conditions, and medication use, may affect physical function capacity among these older adults. These variables were, therefore, not controlled and may possibly, confound the variables studied. Thus, there is the need to control many of these confounding variables in an attempt to explore factors influencing physical function capacity of older adults in future studies.

## Practical Implications

Increased in body weight, poor cardio-respiratory fitness and physical inactivity are key contributors to the decline in physical function ability among these older adults. The study suggests that maintaining a healthy weight and engaging in regular physical activity that promotes cardiovascular endurance can have a positive impact on physical function capacity among these older adults. Encouraging physical activity can help maintain cardiovascular endurance and healthy body weight, which in turn improve physical function ability in these older adults. Health promotion programmes can be designed to increase physical activity levels among these older adults. Also, weight management strategies such as dietary changes and regular physical exercise could be recommended to older adults to maintain a healthy weight.

Given the importance of cardiovascular endurance to physical function ability, older adults should be encouraged to engage in regular physical activity to maintain their cardiovascular health. Furthermore, for older adults who are experiencing physical limitations due to poor cardiovascular endurance or obesity, rehabilitation programmes can be designed to help improve their physical function ability. These programmes can include exercises to increase cardiovascular endurance, muscular endurance and stabilization, flexibility and other activities to improve overall physical function capacity. Finally, the findings can inform health policies aimed at promoting healthy ageing.

## Conclusions

The physical activity levels of older adults in Navrongo were generally low, with accompanied high levels of BMI values, low cardio-respiratory fitness and a decline in physical functional capacity especially among the elderly women. This means that the older adult women in Navrongo may suffer serious health consequences as they are predisposed to non-communicable diseases and other illnesses. Moreover, physical function capacity of these older adults is dependent on their body composition and cardio-respiratory endurance. Thus, increasing the likelihood of increased dependency ratio, with the attendant economic and social challenges. Therefore, evidence-based interventions that promote healthy weight management and cardiovascular fitness may help these adults to maintain their physical independence and improve overall quality of life. Future research could explore the relationship between these factors and physical function ability and identify additional factors that may impact physical function among older adults longitudinally.

## Declarations

### Ethics approval and consent to participate

Ethical clearance and approval were sought from University of Cape Coast Institutional Review Board (ID-UCCIRB/CES/2022/30) for this study. All participants who consented to take part in this study duly signed or thumb printed a consent form provided to them.

### Consent for Publication

Informed consent was obtained from all subjects involved in the study. Participants and the heads of households were duly informed and they gave consent and approval that results and findings from this study could be published.

### Data Availability Statement

The datasets generated and/or analysed during the current study are available in the Open Science Framework, and it is here: https://osf.io/zvdb2

### Author Contributions

Conceptualization and study design: EAA and EWA, Methodology: EAA, EWA, EOS, and MS, Data collection: EAA, EWA, EOS, DA, and SRS, Data analysis: EAA and EWA, Writing - initial manuscript: EAA, Writing - final manuscript: EAA and EWA (with critical feedback provided by EOS, MS, and SRS and DA), Editing and review - All authors edited and considerably reviewed the manuscript, proofread for intellectual content and consented to its publication.

## Acknowledgements

The authors thank the people of Navrongo for their cooperation.

